# Large-scale genome-wide analyses with proteomics integration reveal novel loci and biological insights into frailty

**DOI:** 10.1101/2024.08.26.24312584

**Authors:** Jonathan K.L. Mak, Chenxi Qin, Anna Kuukka, FinnGen, Sara Hägg, Jake Lin, Juulia Jylhävä

## Abstract

Frailty is a clinically relevant phenotype with significant gaps in our understanding of its etiology. We performed a genome-wide association study of frailty in FinnGen (N=500,737) and replicated the signals in the UK Biobank (N=429,463) using polygenic risk scores (PRSs). We prioritized genes through proteomics integration (N∼45,000; UK Biobank) and colocalization of protein quantitative trait loci. Frailty was measured using the Hospital Frailty Risk Score (HFRS). We observed 1,588 variants associated with frailty (*p*<5×10^-8^) of which 1,242 were novel, i.e., previously unreported for any trait. The associations mapped to 106 genes of which 31 were novel. PRS replication validated the signals (β=0.074, *p*<2×10^-16^). Cell type enrichment analysis indicated expression in neuronal cells. Protein levels of *KHK*, *CGREF1*, *MET*, *ATXN2*, *ALDH2*, *NECTIN2*, *APOC1*, *APOE* and *FOSB* were associated with HFRS, whereas colocalized signals were observed within *APOE* and *BRAP*. Our results reveal novel genetic contributions and causal candidate genes for frailty.

---

Aging is a highly complex process with substantial heterogeneity in health trajectories among individuals. Frailty represents a clinically relevant aging phenotype that gauges health in aging^1^ and predicts various adverse outcomes independent of chronological age^2^. Frailty describes a syndrome of decreased physiological reserves across multiple homeostatic systems^1^. Currently, no gold standard exists to measure frailty; instead, several scales with different properties have been developed, each capturing partially different at-risk populations^3^. Created based on 109 weighted International Classification of Diseases, 10^th^ Revision (ICD-10) codes characterizing older adults with high resource use and diagnoses associated with frailty, the Hospital Frailty Risk Score (HFRS) presents a relatively new scale to measure frailty^4^. It has a fair overlap with existing frailty definitions based on the deficit accumulation (frailty index [FI]) and phenotypic (frailty phenotype [FP]) models of frailty and has a moderate agreement with the FI^4^.

The etiology of frailty remains incompletely understood. Twin studies by us and others suggest that frailty, measured using the FI, is up to 52% heritable^5,6^, with relatively stable genetic influences across age^7^. To date, only two previous large-scale genome-wide association studies (GWASs) of frailty exist. Atkins et al. performed a meta-analysis GWAS of FI identified 34 loci and estimated the single nucleotide polymorphism (SNP) heritability of the FI at 11%^8^. Ye at al. identified 123 loci for FP and estimated the SNP heritability of the FP at 6%^9^. It is however likely that additional genetic signals exist and analyses in other large populations can shed further light on the genetic underpinnings of frailty.

To the best of our knowledge, no previous studies into the genetics of frailty using HFRS as the definition exist. To this end, we set out to perform a GWAS of the HFRS in the FinnGen sample (N=500,737), with replication of the signals in using a polygenic risk score of the HFRS in the UK Biobank (N=429,463). As dementia has the highest weight in the HFRS definition, we performed a sensitivity the analysis by removing the contribution of dementia from the HFRS. A functional follow up to identify causal genetic loci was performed through integration of measured protein levels in the UK Biobank (N up to 44,678) and a colocalization analysis of protein quantitative trait locus (pQTL)^10^.

## Results

### Sample characteristics

The workflow of the analyses is presented in **Figure 1**. In the HFRS GWAS and subsequent PRS analyses, we included 500,737 (282,202 females, 56.4%) FinnGen participants and 429,463 UK Biobank participants (232,380 females, 54.1%) of European descent (white British). Characteristics of the study populations are presented in **Table 1**.

**Figure 1.**
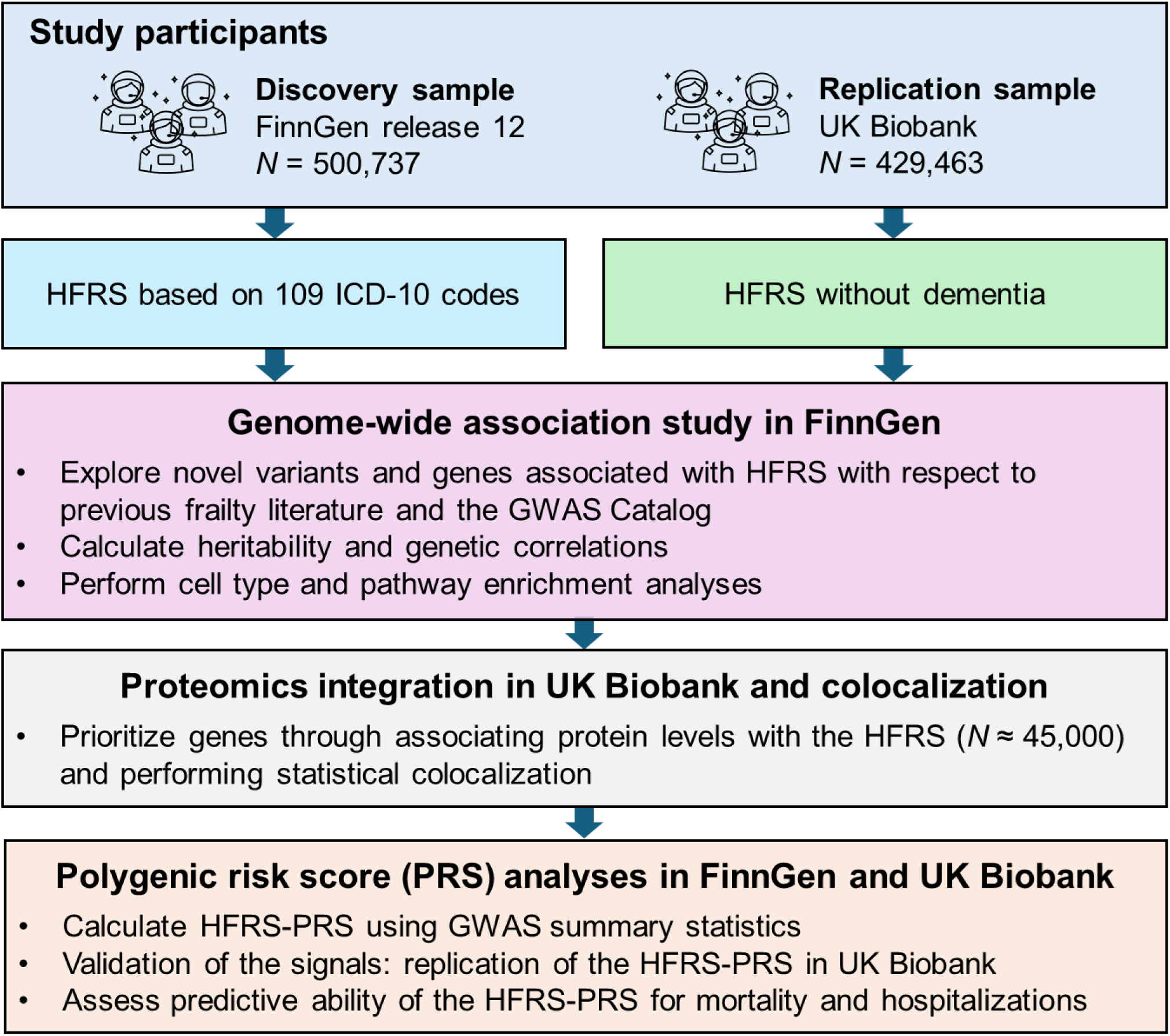
Outline of the study. GWAS, genome-wide association study; HFRS, Hospital Frailty Risk Score; ICD-10, International Classification of Diseases, 10^th^ Revision; PRS, polygenic risk score

**Table 1.** Characteristics of the study samples.

| **Characteristic** | **FinnGen** | **UK Biobank** |
| --- | --- | --- |
| No. of individuals | 519,200 | 429,463 |
| Age at baseline assessment, mean (SD) | 53.1 (17.9) | 56.9 (8.0) |
| Age at end of follow-up/death, mean (SD) | 60.8 (18.0) | 70.8 (7.9) |
| Sex, n (%) |  |  |
| Women | 292,784 (56.4) | 232,380 (54.1) |
| Men | 226,416 (43.6) | 197,083 (45.9) |
| BMI (kg/m<sup>2</sup>), mean (SD) | 27.35 (5.53) | 27.41 (4.76) |
| Missing, n (%) | 142,454 (27.4) | 1348 (0.3) |
| Smoking, n (%) |  |  |
| Non-smoker | 156,355 (50.9) | 232,968 (54.2) |
| Former smoker | 70,317 (22.9) | 151,248 (35.2) |
| Current smoker | 80,736 (26.2) | 43,776 (10.2) |
| Missing | 211,792 | 1,471 |
| HFRS, median (IQR) | 5.2 (1.6–10.4) | 1.5 (0–5) |
| Women, median (IQR) | 5.3 (1.6–10.5) | 1.5 (0–4.7) |
| Men, median (IQR) | 5.0 (1.5–10.3) | 1.5 (0–5.4) |
| HFRS categories, n (%) |  |  |
| Low risk (<5) | 241,656 (48.4) | 320,961 (74.7) |
| Intermediate risk (5–15) | 188,147 (37.8) | 78,292 (18.2) |
| High risk (>15) | 65,925 (13.2) | 30,210 (7.0) |
| HFRS >5, n (%) | 254,874 (51.0) | 106,645 (24.8) |
| HFRS >5 before age 65, n (%) | 95,410 (18.4) | 35,556 (8.3) |
| Died during follow-up, n (%) | 62,764 (12.1) | 38,636 (9.0) |
| Time to mortality follow-up (year), median (IQR) | 4.4 (2.6–8.5) | 14.4 (13.6–15.0) |
| Number of hospitalizations, median (IQR) | 8 (4–17) | 1 (0–3) |

Note. FinnGen participant characteristics are presented for the sample with non-missing phenotypic data (N=519,200).
BMI, body mass index; HFRS, Hospital Frailty Risk Score; IQR, interquartile range; SD, standard deviation.

### GWAS of HFRS

We identified 1,588 variants associated (*p*<5×10^-8^) with the HFRS in the main analysis and 492 variants in the sensitivity analysis removing the dementia weights from the HFRS (**Figure 2a** & **b**; **Supplementary Tables 1** & **2**). As dementia diagnosis has the highest weight in the HFRS formula, the most influential peak expectedly resided in the *APOE* (rs7412) region on chromosome 19 (**Figure 2a**). Sensitivity analysis confirmed the expected loss of the *APOE* peak (**Figure 2b**). Of the 1,588 and 492 variants associated with HFRS and HFRS without dementia, 1,242 and 440, respectively, were novel with respect to the GWAS Catalog and previously reported GWAS results on the FI^8^, FP^9^ and mvAge^11^ (**Supplementary Tables 1** & **2**). The variants mapped to 106 and 50 genes of which 31 and 8 were novel, i.e., previously unreported for any trait at *p*<5×10^-8^, also revealing unique (non-shared) associations in both analyses (**Figure 3a**, **Supplementary Tables 1** & **2**). The overlap between our findings and previous GWAS on frailty and mvAge is presented individually for each GWAS gene set in **Supplementary Figure 1**.

**Figure 2.**
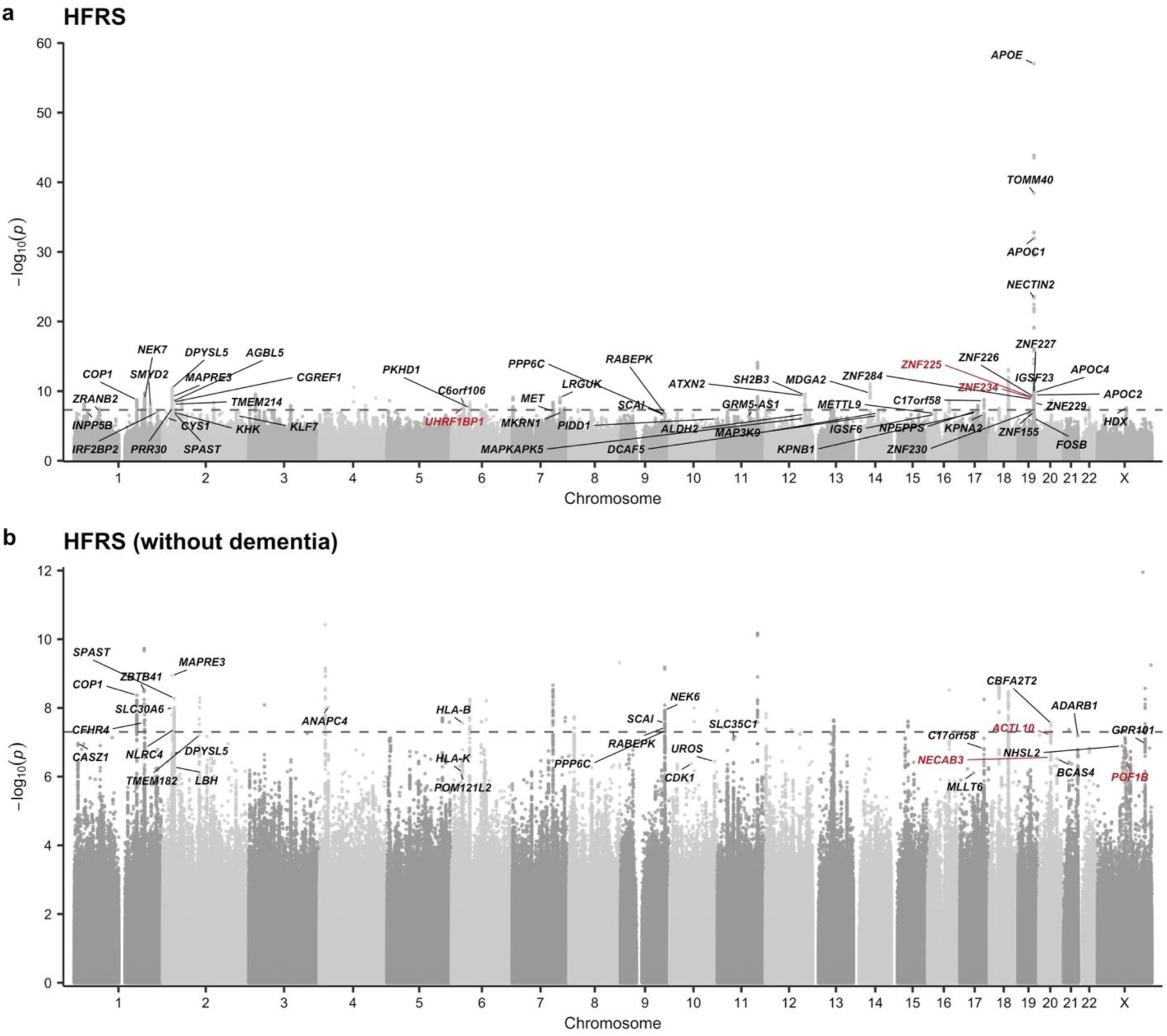
Manhattan plots for the associations with **(a)** Hospital Frailty Risk Score (HFRS) and **(b)** HFRS excluding dementia in FinnGen. The dashed lines indicate the genome-wide significance threshold (*p*=5×10^-8^). The annotations represent the strongest signals in genes containing potentially functional variants (*p*<5×10^-7^) associated with frailty; red font indicates genes that include variants previously unreported in the GWAS Catalog or previous GWASs of frailty.

**Figure 3.**
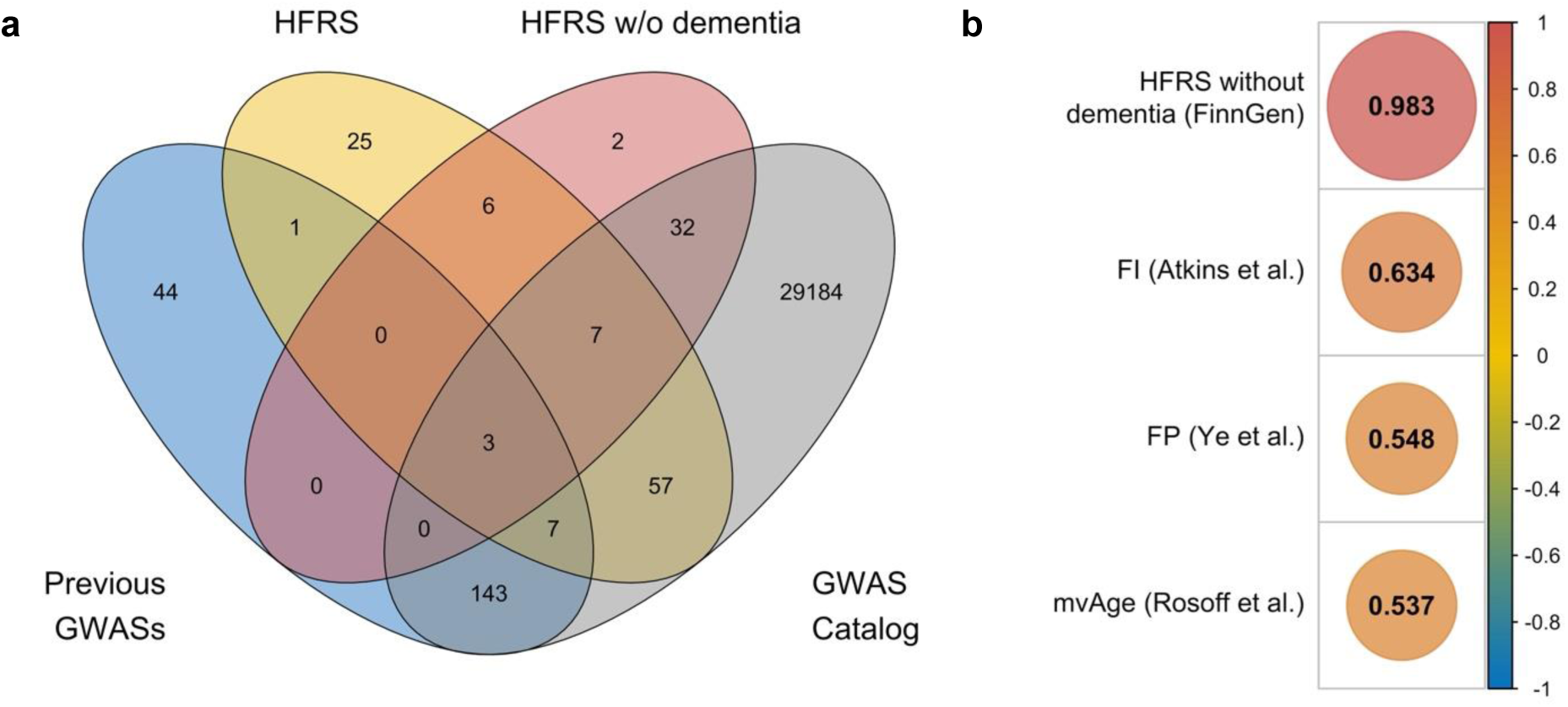
Novel genes and genetic correlations of the with other related traits. **(a)** Venn diagram showing the overlap of genes associated with the full HFRS and the HFRS without dementia at *p*<5×10^-8^ in FinnGen and those reported in the literature. Previous GWASs refers to genes identified in for the FI (Atkins et al., 2021), FP (Ye et al., 2023), and mvAge (Rosoff et al., 2023). **(b)** Genetic correlations between HFRS in FinnGen and other frailty-related traits. All the correlations were statistically significant at *p*<2.2×10^-16^. FI, frailty index, FP, frailty phenotype; GWAS, genome-wide association study; HFRS, Hospital Frailty Risk Score; PRS, polygenic risk score

### Genetic correlation and heritability

We observed a lambda genomic control value of 1.27 with an intercept of 1.19 (standard error [SE]=0.011) for HFRS and 1.11 with an intercept of 1.23 (SE=0.010) for HFRS without dementia (QQ plots provided in **Supplementary Figure 2**). Despite the relatively high lambda values, the intercepts suggest that the inflation in test statistics was mainly due to polygenicity, rather than bias due to population stratification. The single nucleotide variant (SNP) heritability was 0.06 (SE=0.002) for HFRS and 0.04 (SE=0.002) for HFRS without dementia. Statistically significant and positive genetic correlations (*p*<2.2×10^-16^) were observed between HFRS and previous GWASs on frailty and mvAge (**Figure 3b)**.

### Cell type and pathway enrichment

For HFRS, the top (*p*<3.7×10^-5^, corrected for multiple testing) cell types enriched for expression were limbic system neurons in cerebrum, excitatory neurons (Ex6) in visual cortex, oligodendrocyte precursor cells (OPCs) in cerebellar hemisphere and oligodendrocytes in cerebellum (**Supplementary Figure 3** & **Supplementary Table 3**). For HFRS without dementia, the top cell types were OPCs and astrocytes in cerebellar hemisphere, skeletal muscle satellite cells in muscle, endocrine cells in stromal cells in stomach (**Supplementary Figure 4** & **Supplementary Table 4**). Enrichr^12^ pathway analysis (adjusted *p*<0.05) showed that the top pathways for HFRS functions relevant to the nervous system (Herpes simplex virus 1 infection, Netrin Mediated Repulsion Signals), cell adhesion and lipid metabolism (**Supplementary Table 5**). For HFRS without dementia, none of the pathways were significant after multiple testing correction (**Supplementary Table 6**).

### Exploring potentially causal and functional variants through proteomics integration

To identify potentially causal and functional variants (i.e., missense, splice region, loss of function and 5’ and 3’ untranslated region [UTR] variants associated with the HFRS and HFRS without dementia at *p*<5×10^-7^) (**Supplementary Tables 7–8**), we associated the protein levels of the corresponding prioritized genes to HFRS (13 proteins available in UK Biobank Olink platform) and HFRS without dementia (8 proteins available in UK Biobank Olink platform). After adjusting for birth year, sex, and the first 10 principal components (PCs), 9/13 (KHK, CGREF1, MET, ATXN2, ALDH2, NECTIN2, APOC1, APOE and FOSB) and 2/8 (CDK and POF1B) proteins were significantly associated with the HFRS and HFRS without dementia, respectively, at a false discovery rate (FDR) <0.05 (**Figure 4** & **Supplementary Table 9**).

**Figure 4.**
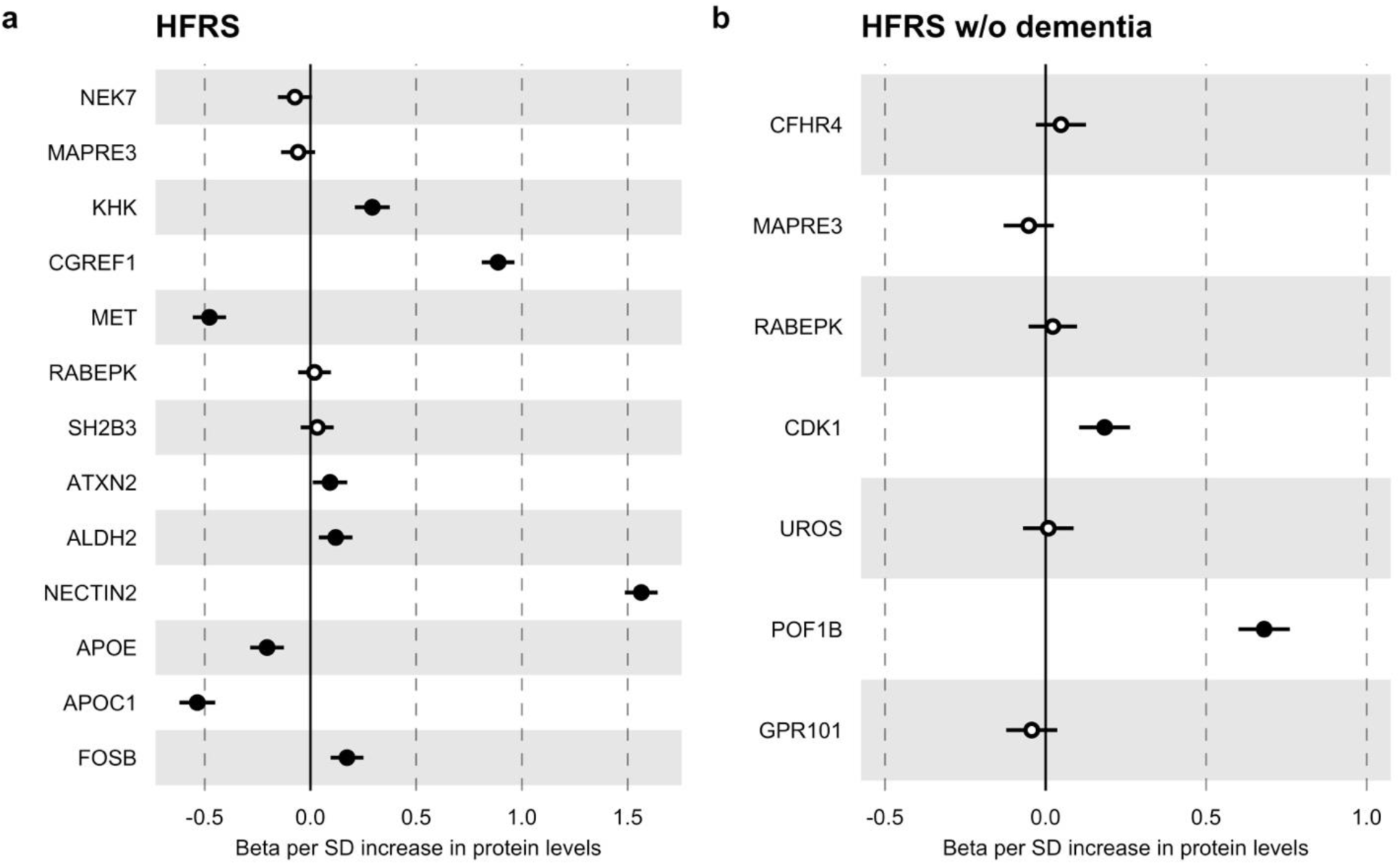
Protein associations with the **(a)** full HFRS and **(b)** HFRS without dementia the in UK Biobank using linear regression models. All models were adjusted for birth year, sex, and the first 10 principal components. Solid dots indicate significant associations at a false discovery rate <0.05. HFRS, Hospital Frailty Risk Score; SD, standard deviation

### Colocalization analysis

We further conducted pQTL colocalization analyses for the 24 loci identified for HFRS and 15 loci identified for HFRS without dementia GWASs (**Supplementary Tables 10and 11**). A total of 20 loci for HFRS and 9 loci for HFRS without dementia had enough power for the analyses (posterior probability > 0.88, see Methods). Of them, the colocalized signal (i.e., shared single causal variant, H4<90, see Methods) was detected within *APOE* and *BRAP* genes for HFRS (**Supplementary Table 10**), whereas no colocalized signal was detected within genes for HFRS without dementia. For most of the tested genes, the H3 values were greater than or close to 90, indicative of distinct causal variants for protein levels and HFRS (**Supplementary Tables 10and 11**). Regional association plots of the *APOE* gene demonstrated that the strongest signal peak rs429358 and variants in high LD with it fall in the vicinity (**Supplementary Figure 5**A).

*HFRS PRS analyses in FinnGen and UK Biobank: early-onset frailty and outcome prediction* The PRS of the HFRS (PRS-HFRS) was associated with HFRS in the full sample of the UK Biobank (β=0.074 per SD increase; *p*<2×10^-16^) after adjusting for birth year, sex, smoking and first 10 PCs (**Figure 5a**). Next, using similar adjustments, we analyzed whether the HFRS-PRS could predict early-onset frailty i.e., HFRS>5 before age 65, and observed an odds ratio of 1.25 (*p*<2×10^-16^) in the sample of all self-identified whites of the UK Biobank (**Figure 5b**). The estimates of the HFRS-PRS were essentially similar in men and women compared to the full sample across all analyses (**Figure 5a–d**). The numeric results of all the HFRS-PRS analyses are presented in **Supplementary Table 14**. Lastly, we examined whether the HFRS-PRS predicts all-cause mortality and number of hospitalizations and found significant associations with both outcomes (**Figure 5c and d**); adjusting for the HFRS-PRS based on a crude model with age and sex improved model performances (**Supplementary Table 11**).

**Figure 5.**
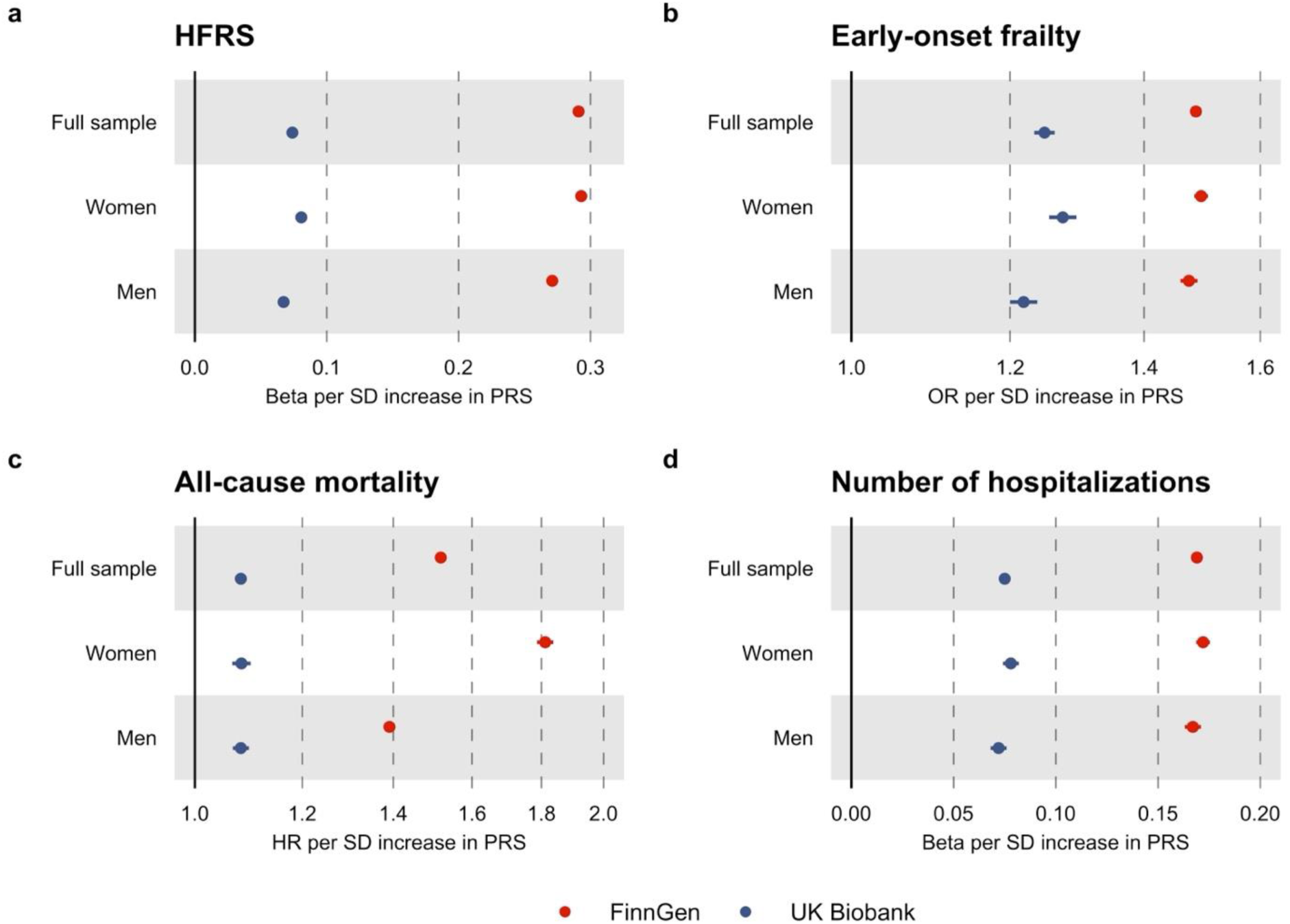
Associations of the. HFRS-PRS with the HFRS (a), early-onset frailty (b), all-cause mortality and number of hospitalizations (d) in FinnGen and UK Biobank. All models included birth year, birth region, sex, smoking and first 10 principal components as covariates. HFRS, Hospital Frailty Risk Score; HR, hazard ratio; OR, odds ratio; PRS, polygenic risk score; SD, standard deviation.

## Discussion

Our results represent the largest GWAS of frailty to date and the first GWAS of frailty assessed through the HFRS, revealing 1,588 variants, of which 1,242 were novel i.e., previously unreported for any trait. The variants mapped to 106 genes, of which 31 were novel and highlights that the genetic etiology of frailty is largely unrelated to previously known disease risk variants. Protein levels of *KHK*, *CGREF1*, *MET*, *ATXN2*, *ALDH2*, *NECTIN2*, *APOC1*, *APOE* and *FOSB* were associated with HFRS, whereas colocalized signals were observed within *APOE* and *BRAP*. Enriched expression of the associated genes was observed in various neuronal cells, also when the contribution of dementia was removed from the frailty definition. Using the HFRS-PRS, we replicated the genetic signals in an independent sample (UK Biobank) and validated our findings (β=0.074, *p*<2×10^-16^). The HFRS-PRS also predicted early-onset frailty as well as all-cause mortality and number of hospitalizations.

The strongest GWAS signals were observed in the *TOMM40*/*APOE*/*APOC1*/*NECTIN2* locus on 19q13.3, a locus in strong LD and known for its associations with cognitive^13^ and cardiometabolic^14^ traits. We observed the strongest signal for the missense variant rs429358 (388 T > C) that together with rs7412 defines the *APOE* ε2, ε3, and ε4 haplotypes. The rs7412 was however not associated with frailty in our study. A similar pattern of finding has been observed for longitudinal weight loss – a feature that also characterizes frailty – where rs429358 increased the risk, while rs7412 did not^15^. Previous studies have shown that this locus is pleiotropic, such that rs429358 influences cognitive traits, while rs7412 controls plasma lipid levels^16^. We did nevertheless identify lipid-level-increasing variants, such as the *APOC1* rs4420638 (G allele)^17^ associated with frailty, but lipid-associated variants were not abundant in our signals. Our sensitivity analysis removing the contribution of dementia from the HFRS truncated the chromosome 19 peak as expected and revealed additional loci. Of the 106 genes identified for HFRS, 16 were shared with HFRS without dementia, while 34 genes were unique to HFRS without dementia. Genetic correlation between HFRS and HFRS without dementia was nevertheless almost perfect (0.98), indicating the same underlying genetic construct.

Intersecting the HFRS-associated signals with previous frailty GWASs of FI^8^ and FP^9^ revealed a negligible overlap. Genetic correlations between HFRS, FI and FP were nevertheless moderate, ranging from 0.54 to 0.63. We estimated the SNP heritability of HFRS at 6%, an estimate in the same range as previously reported for the FI (11%)^8^ and FP (6%)^9^. In our previous study^18^, we assessed the phenotypic correlation between HFRS and FI at 0.21 and HFRS and FP at 0.31 in the UK Biobank participants, indicating somewhat lower than phenotypic correlations compared to their genotypic counterparts. These findings thus suggest that while the different operationalizations of frailty share their genetic etiologies to a significant extent, environmental risk factors and relevant interactions contributing to the expression of frailty may differ.

Cell type enrichment indicated enriched expression of the genes associated with the signals in various neuronal cells, such as limbic system and excitatory neurons, OPCs and oligodendrocytes located in the cerebrum, visual cortex, cerebellar hemisphere and cerebellum, respectively. Enrichment of OPCs (cerebellar hemisphere) persisted also after removing the contribution of dementia diagnoses from the HFRS. Expression enrichment in brain tissues was likewise observed in our previous GWAS of FI^8^ in which we identified frontal cortex BA9, cerebellar hemisphere, spinal cord cervical c-1 and hippocampus as significant. The GWAS on FP^9^ by Ye et al. also identified their genetic signals enriched in brain tissues, such as cerebellar hemisphere, frontal cortex BA9 and cerebellum. What is noteworthy is that neither FI or FP include any items of cognition or dementia diagnosis in the frailty definition. Our findings thus reinforce the role of central nervous system functions in frailty, regardless of the definition. The previous FI and FP GWAS signals were also enriched in inflammatory mechanisms or pathways^8,9^, a finding not observed by us with the exception of the Herpes simplex virus 1 infection pathway. Our pathway analyses instead highlighted cell adhesion and lipid metabolism relevant to the signals. Our results included several cell adhesion molecules, such as *CNTNAP2*, *CADM1*, *NCAM1*, *PVR*, *NECTIN2*, suggesting novel contributions to frailty. While cell adhesion molecules mediate the transport of leukocyte migration towards the inflammation site^19^, previous results linking cell adhesion directly to frailty are scarce, except for the association of circulating ICAM-1 with frailty^20^.

The protein level associations of the potentially functional variants with frailty revealed the largest effect sizes for CGREF1, NECTIN2, MET and APOC1, with elevated levels of the former two and lower levels of the latter two associating with higher HFRS score. CGREF1 is a secretory cell growth regulator whose involvement in disease is currently unknown. A previous GWAS has however demonstrated associations of *CGREF* variants with plasma lipids^21^. *NECTIN2*, a cell adhesion molecule and mediator of viral entry into neuronal cells has been linked to Alzheimer’s disease^22^ and plasma lipid profiles^21^ in previous GWASs. Elevated serum levels of NECTIN2 have been reported in colorectal cancer^23^. *MET* is a proto-oncogene and a receptor tyrosine kinase with previous GWAS findings on body height and liver enzymes^24^ but limited evidence on genetic disease associations. We found that lower plasma levels of APOC1 and APOE were associated with greater frailty, a finding that is in line with previous results on low APOE levels associated with progression of cognitive impairment^25^ and dementia-related mortality^26^. Findings on all-cause, cardiovascular and cancer mortality and APOE^26^, and hyperlipidemia and APOC1^27^, nevertheless demonstrate higher levels of these proteins associated with increased risks, indicating pleiotropic functions of these proteins. When the dementia weights were removed from the HFRS, higher plasma levels of CDK1 and POF1B were associated with greater frailty. Previous findings on variants in these genes are limited to height^28^ and bone mineral density^29^ for *CDK1* and velopharyngeal dysfunction^30^ for *POF1B*. Results from the pQTL colocalization analysis suggest that the same causal variants in *APOE* and *BRAP*, a BRCA1 associated protein, underlie the protein level and HFRS. Most tested genes nevertheless showed distinct causal variants for the proteins and HFRS.

Replication of the GWAS signals through the HFRS-PRS in the UK Biobank validated the results, including individually for men and women. We also showed that the HFRS-PRS can identify individuals at risk of early-onset frailty. As frailty manifests relatively late in life for most individuals, risk assessment through PRS may offer possibilities for early intervention to mitigate frailty before it escalates where prevention is still effective. PRSs of various age-related phenotypes associated with negative outcomes, such as frailty, epigenetic clocks and functional capacity could perhaps be jointly considered to yield more robust predictions. Future studies are however needed to ascertain the clinical utility of such approaches.

This study has several strengths, the most notable being the large sample size, equaling to ∼1 million participants. Functional follow-up through proteomics integration provided additional insight into the roles of the identified genes in frailty. Our definition of frailty was based on clinical diagnoses in register data; such an approach has both advantages and disadvantages. A notable advantage is that in Finland and the UK, public healthcare is primarily tax-funded, and each citizen has equal access. With a diagnosis-based ascertainment of frailty, issues pertinent to self-reported data, such as recall bias and missing information were avoided. On the other hand, some conditions may be underreported in the registers, while others may have a lag from the onset of symptoms to assigning the diagnosis. We also note that the HFRS-PRS associations were weaker in the UK Biobank compared to FinnGen, a finding likely explained by healthy selection due to volunteer-based participation to the UK Biobank compared to FinnGen that consists of national cohorts and biobank samples of hospitalized individuals. Also pertinent to all GWASs, the discovery samples tend to have stronger association statistics compared to replication, a phenomenon known as the winner’s curse.

In conclusion, our results provide the first GWAS on HFRS and reveal novel genetic contributions and causal candidate genes. Our results also highlight previously unreported associations between cell adhesion molecules and frailty. Overall, the results reinforce previous findings that central nervous functions are relevant to the etiology of frailty, regardless of how frailty is defined.

## Methods

### Samples

FinnGen is a large national genetic resource (N=520,210; Release 12) established in 2017 and consisting of Finnish individuals, aged 18 years and older at study baseline^31^. FinnGen includes prospective epidemiological and disease-based cohorts as well as hospital biobank samples. Information on diagnoses since 1969 was linked by the unique national personal identification number to national healthcare, population and cause of death registries and recorded using the ICD Revisions 8–10. Information on dates and causes of death were obtained via linkages to the population and cause of death registers through (September 30, 2023, R12 v1). After excluding individuals with missing information on baseline age, birth year and sex, and samples not passing genotyping quality control (see below), we included 500,737 FinnGen participants in this study.

The UK Biobank includes 502,642 volunteer participants, aged 37 to 73 years old at baseline, recruited through 22 assessment centers across England, Scotland and Wales between 2006 and 2010^32^. The participants provided self-reported information on demographics, lifestyle and disease history via questionnaire and underwent physiological measurements, including providing a blood sample for genetics data. Hospital inpatient data were sourced from the Hospital Episode Statistics containing electronic medical records (i.e., ICD-10 codes) for all hospital admissions to National Health Service hospitals in England through December 31, 2022. Death register data covered all deaths in the population through December 31, 2022, including primary and contributory causes of death. Ethics statements of FinnGen and UK Biobank are presented in Supplementary methods.

### Assessment of frailty

The HFRS was calculated according to a previously described protocol^4^ based on 109 weighted ICD-10 codes, such that each code was assigned with a weight ranging from 0.1 to 7.1 according to the strength of the association with frailty (**Supplementary Table 12**). The HFRS score was then calculated by summing up all the weights and used as a continuous variable in the GWAS. We also categorized the HFRS into low (<5), intermediate (5–15) and high (>15) risk of frailty as previously described^4^ and used the cut points to describe frailty in our study populations. In the main analysis, we included all available ICD-10 codes for each person from age 30 years to the age at the end of follow-up to calculate the HFRS. As dementia diagnoses have the highest weight tin the HFRS, we calculated the HFRS also by excluding dementia weights form the formula and performed all analyses, except for PRS associations, using the HFRS without dementia.

### Genotyping and imputation

Genotyping in FinnGen was performed on Illumina (Illumina Inc., San Diego, CA) and custom AxiomGT1 Affymetrix (Thermo Fisher Scientific, Santa Clara, CA) genome-wide arrays and imputed to 16,387,711 (INFO > 0.6) variants using a population-specific SISu v.3 imputation reference panel as previously described^33^. Individuals with ambiguous sex and non-Finnish ancestry were excluded. UK Biobank samples (v3 genotyping release) were genotyped on custom Affymetrix microarrays and imputed using the 1000 Genomes and the Haplotype Reference Consortium reference panels to ∼93M variants^34^. Participants were excluded if they were flagged as having unusually high heterozygosity or missing genotype calls (<5%). Our analysis was restricted to white British participants (N=429,463). Detailed procedures on genotype calling, quality controls and imputation have been previously described for FinnGen^31^ and UK Biobank^34^.

### GWAS

The analytical pipeline for GWAS and post-GWAS analyses is presented in **Figure 1**. We first performed a GWASs of HFRS in FinnGen using the SAIGE^35^ (v0.35.8.8) software, which uses linear mixed-effects modeling to account for genetic relatedness and confounding by ancestry^36^. We included variants (N=21,294,561) with minor allele frequency >0.01%, Hardy-Weinberg *p*- value >1×10^-9^ and imputation INFO score ≥0.9. The models were adjusted for birth year, birth region, sex and the 10 first PCs. HFRS was inverse normal transformed prior to modeling. The genome-wide significance level was set to 5×10^-8^. Using the GWAS Catalog and results of previous GWASs into frailty (using the FP^9^ and FI^8^ to measure frailty) and mvAge^11^, a genomic structural equation modeling-derived composite construct of healthspan, parental lifespan, extreme longevity, frailty and epigenetic aging, we assessed the number of novel and previously unreported associations.

### Genetic correlation and heritability

Using linkage disequilibrium score regression^37^ (v1.0.1) and LD merged with the HapMap3 reference panel of ∼1.1 million variants, we estimated 1) the potential bias from e.g. population stratification and cryptic heritability in the GWAS results, 2) heritability of HFRS and 3) genetic correlations between HFRS and previous GWASs of FI^8^, FP^9^ and mvAge^11^. As the FI GWAS^8^ used an opposite effect allele compared to the standard FinnGen workflow, we inverted the genetic correlation coefficient to facilitate interpretation.

### Functional annotation: cell type and pathway enrichment

To explore tissue and cell type specificity of the annotated genes underlying HFRS, we applied WebCSEA, a web platform to derive context-specific expression patterns of genes underlying complex traits, encompassing the Human Cell Atlas and single cell data resources^38^. Enrichr pathway analysis^12^ based on KEGG^39^, Reactome^40^ and WikiPathway^41^ resources, was applied to explore enriched pathways (FDR<0.05) of the identified genes (GWAS *p*<5×10^-8^).

### Proteomics integration

To prioritize genes and identify potentially functional and causal variants, we narrowed down the association signals to a smaller number of missense, splice region, loss of function and 5’ and 3’ UTR variants (the two last mentioned potentially affecting transcript stability, localization and signal response) identified from the Variant Effect Predictor pipeline^42^, that were associated with the HFRS at a slightly more relaxed threshold (*p*<5×10^-7^). Using the Olink proteomics data, we then examined if the protein levels of the variants (at a gene level resolution) were associated with HFRS in the UK Biobank. Details of the UK Biobank Olink proteomics assay, quality control and data processing procedures have been described elsewhere^43^. Briefly, ∼50,000 UK Biobank participants were randomly selected for the proteomics profiling using EDTA plasma samples collected at the baseline assessment. A total of 2,923 proteins was measured across 8 protein panels using the antibody-based Olink Explore 3072 platform. Protein levels were measured in normalized Protein eXpression (NPX) values, which represent the relative concentration of proteins on a log- 2 scale. All the protein levels were scaled to mean=0 and SD=1 before the association testing. Linear regression models were then performed to assess the association between the proteins that were available in the Olink platform and HFRS, adjusting for birth year, sex, and the first 10 PCs. We considered an FDR<0.05 as statistically significant in the proteomics analysis.

### Colocalization analyses

To further prioritize the genes, we summarized gene loci to which the genome-wide significant or potential functional variants were mapped (**Supplementary Tables 1, 2, 7 & 8**). We performed a Bayesian-based colocalization analysis for each locus, using a flanking window of 1Mb and default parameters for prior probabilities^10^. The analysis assumes that only one causal variant exists for each trait in a genomic locus and returns posterior probabilities indicating the likelihood that the following hypotheses (H) are true: there is no association at the locus with either protein level or HFRS (H0); there is an association with protein level but not HFRS (H1); there is no association with protein level but there is an association with HFRS (H2); there is an association with both the protein level and HFRS but with distinct causal variants (H3); there is an association with both the protein level and HFRS with a shared causal variant (H4). We considered the analysis having enough power if the sum posterior probabilities of having a distinct or shared causal variant exceeded 88%. A colocalized signal was detected if the posterior probability of a shared causal variant (H4) existence was greater than 90%.

### PRS analyses

Using the GWAS summary statistics from FinnGen, we calculated the PRS for HFRS by applying PRS with continuous shrinkage^44^ (PRS-CS) and using the European panel from the 1000 Genomes^45^ LD reference, where ∼1.1 million variants were selected. All the PRS analyses were performed in FinnGen (the discovery sample) for reference and replicated in the UK Biobank. Using linear regression, we fitted linear model to assess how the HFRS-PRS associates with the HFRS. HFRS was considered as a standardized z-score in the linear regressions. We also performed logistic regressions to assess the associations of the HFRS-PRS with early-onset frailty, defined as HFRS >5 before age 65. The PRS was modeled as per SD change and all the models included birth year, birth region (FinnGen), sex and the first 10 PCs as covariates.

Lastly, as frailty manifests in late life for most individuals, we asked whether the HFRS- PRSs could be used in early risk stratification to identify individuals at risk of adverse outcomes. To this end, Cox models with attained age as the timescale and linear regression models were fitted to assess whether the HFRS-PRS predicts all-cause mortality and number of hospitalizations, respectively. The added value of the HFRS-PRS beyond age and sex in the prediction was assessed using the F-test for linear regressions and likelihood ratio test for Cox models. The number of hospitalizations was scaled to a mean=0 and SD=1 prior to modeling.

## Data availability

Individual-level data cannot be stored in public repositories or otherwise made publicly available due to ethical and data protection restrictions. However, data are available upon request for researchers who meet the criteria for access to confidential data. Data from the UK Biobank are available to bona fide researchers upon application at https://www.ukbiobank.ac.uk/enable-your-research. FinnGen results, according to FinnGen consortium agreement, are subjected to one year embargo and summary statistics are then made available to the scientific community and release two times a year. Information on accessing FinnGen data can be found at https://www.finngen.fi/en/access_results.

## Code availability

All the data processing, visualization, and statistical analyses were performed using Python 3.8 (2.7 for LDSC) and R v.4.3.2 (R Foundation for Statistical Computing, Vienna, Austria; https://www.r-project.org/). Venn diagrams were created using the R package *ggvenn* (version 0.1.10; https://cran.r-project.org/web/packages/ggvenn/index.html). Correlation plots were created using the R package corrplot (v.0.92; https://cran.r-project.org/web/packages/corrplot/index.html). Forest plots were created using the R package *ggforestplot* (v.0.1.0; https://nightingalehealth.github.io/ggforestplot/). R codes used to create the figures are available from the authors upon request.

## Acknowledgements

This work was supported by the Swedish Research Council (grant no. 2018-02077 to JJ, 2019- 01272, 2020-06101, 2022-01608), the Research Council of Finland to JJ (grant no. 3493358), the Sigrid Jusélius Foundation to JJ, the Yrjö Jahnsson Foundation to JJ (grant no. 20217416), Instrumentarium Science Foundation to JJ and Signe and Ane Gyllenberg Foundation to JJ (grant no. 6226). This research was conducted using the UK Biobank resource, as part of the registered project 22224. The analyses of UK Biobank genotypes were enabled by resources in project sens2017519 provided by the National Academic Infrastructure for Supercomputing in Sweden (NAISS) at UPPMAX, funded by the Swedish Research Council through grant agreement no. 2022-06725. The FinnGen project is funded by two grants from Business Finland (HUS 4685/31/2016 and UH 4386/31/2016) and the following industry partners: AbbVie Inc., AstraZeneca UK Ltd, Biogen MA Inc., Bristol Myers Squibb (and Celgene Corporation & Celgene International II Sàrl), Genentech Inc., Merck Sharp & Dohme Corp, Pfizer Inc., GlaxoSmithKline Intellectual Property Development Ltd., Sanofi US Services Inc., Maze Therapeutics Inc., Janssen Biotech Inc, and Novartis AG. Following biobanks are acknowledged for delivering biobank samples to FinnGen: Auria Biobank (www.auria.fi/biopankki), THL Biobank (www.thl.fi/biobank), Helsinki Biobank (www.helsinginbiopankki.fi), Biobank Borealis of Northern Finland (https://www.ppshp.fi/Tutkimus-ja-opetus/Biopankki/Pages/Biobank-Borealis-briefly-in-English.aspx), Finnish Clinical Biobank Tampere (www.tays.fi/en-US/Research_and_development/Finnish_Clinical_Biobank_Tampere), Biobank of Eastern Finland (www.ita-suomenbiopankki.fi/en), Central Finland Biobank (www.ksshp.fi/fi-FI/Potilaalle/Biopankki), Finnish Red Cross Blood Service Biobank (www.veripalvelu.fi/verenluovutus/biopankkitoiminta) and Terveystalo Biobank (www.terveystalo.com/fi/Yritystietoa/Terveystalo-Biopankki/Biopankki/). All Finnish Biobanks are members of BBMRI.fi infrastructure (www.bbmri.fi). Finnish Biobank Cooperative -FINBB (https://finbb.fi/) is the coordinator of BBMRI-ERIC operations in Finland. The Finnish biobank data can be accessed through the Fingenious^®^ services (https://site.fingenious.fi/en/) managed by FINBB.

## Author contributions

JJ conceived the study plan and designed the proof outline. JKLM, CQ, JL and AK performed the analyses. JJ, JL and SH were responsible of data acquisition. All authors contributed to the writing of the manuscript and interpretation of the results. All authors listed under FinnGen contributed to the generation of the primary data of the FinnGen data release 12. FinnGen authors are listed in the **Supplementary Table 15**.

## References

1. Clegg, A., Young, J., Iliffe, S., Rikkert, M. O. & Rockwood, K. Frailty in elderly people. The Lancet 381, 752–762 (2013).

2. Kojima, G., Iliffe, S. & Walters, K. Frailty index as a predictor of mortality: a systematic review and meta-analysis. Age Ageing 47, 193–200 (2018).

3. Theou, O., Brothers, T. D., Mitnitski, A. & Rockwood, K. Operationalization of Frailty Using Eight Commonly Used Scales and Comparison of Their Ability to Predict All-Cause Mortality. J. Am. Geriatr. Soc. 61, 1537–1551 (2013).

4. Gilbert, T. et al. Development and validation of a Hospital Frailty Risk Score focusing on older people in acute care settings using electronic hospital records: an observational study. Lancet Lond. Engl. 391, 1775–1782 (2018).

5. Young, A. C. M., Glaser, K., Spector, T. D. & Steves, C. J. The Identification of Hereditary and Environmental Determinants of Frailty in a Cohort of UK Twins. Twin Res. Hum. Genet. Off. J. Int. Soc. Twin Stud. 19, 600–609 (2016).

6. Mak, J. K. L. et al. Sex differences in genetic and environmental influences on frailty and its relation to body mass index and education. Aging 13, 16990–17023 (2021).

7. Mak, J. K. L. et al. Genetic and Environmental Influences on Longitudinal Frailty Trajectories From Adulthood into Old Age. J. Gerontol. A. Biol. Sci. Med. Sci. 78, 333–341 (2023).

8. Atkins, J. L. et al. A genome-wide association study of the frailty index highlights brain pathways in ageing. Aging Cell 20, e13459 (2021).

9. Ye, Y. et al. A genome-wide association study of frailty identifies significant genetic correlation with neuropsychiatric, cardiovascular, and inflammation pathways. GeroScience 45, 2511–2523 (2023).

10. Giambartolomeie, C., et al. Bayesian test for colocalisation between pairs of genetic association studies using summary statistics. PLoS 10(5):e1004383 (2014).

11. Rosoff, D. B. et al. Multivariate genome-wide analysis of aging-related traits identifies novel loci and new drug targets for healthy aging. *Nat*. Aging 3, 1020–1035 (2023).

12. Xie, Z. et al. Gene Set Knowledge Discovery with Enrichr. Curr. Protoc. 1, e90 (2021).

13. Aslam, M. M. et al. Genome-wide analysis identifies novel loci influencing plasma apolipoprotein E concentration and Alzheimer’s disease risk. Mol. Psychiatry 28, 4451–4462 (2023).

14. Yeh, K.-H. et al. Genetic Variants at the APOE Locus Predict Cardiometabolic Traits and Metabolic Syndrome: A Taiwan Biobank Study. Genes 13, 1366 (2022).

15. Kemper, K. E. et al. Genetic influence on within-person longitudinal change in anthropometric traits in the UK Biobank. Nat. Commun. 15, 3776 (2024).

16. Bennet, A. M. et al. Pleiotropy in the presence of allelic heterogeneity: alternative genetic models for the influence of APOE on serum LDL, CSF amyloid-β42, and dementia. J. Alzheimers Dis. JAD 22, 129–134 (2010).

17. Willer, C. J. et al. Newly identified loci that influence lipid concentrations and risk of coronary artery disease. Nat. Genet. 40, 161–169 (2008).

18. Mak, J. K. L., Kuja-Halkola, R., Wang, Y., Hägg, S. & Jylhävä, J. Frailty and comorbidity in predicting community COVID-19 mortality in the U.K. Biobank: The effect of sampling. J. Am. Geriatr. Soc. 69, 1128–1139 (2021).

19. Luster, A. D., Alon, R. & von Andrian, U. H. Immune cell migration in inflammation: present and future therapeutic targets. Nat. Immunol. 6, 1182–1190 (2005).

20. Zhang, L., Zeng, X., He, F. & Huang, X. Inflammatory biomarkers of frailty: A review. Exp. Gerontol. 179, 112253 (2023).

21. Sinnott-Armstrong, N. et al. Genetics of 35 blood and urine biomarkers in the UK Biobank. Nat. Genet. 53, 185–194 (2021).

22. Jansen, I. E. et al. Genome-wide meta-analysis identifies new loci and functional pathways influencing Alzheimer’s disease risk. Nat. Genet. 51, 404–413 (2019).

23. Karabulut, M. et al. Serum nectin-2 levels are diagnostic and prognostic in patients with colorectal carcinoma. Clin. Transl. Oncol. Off. Publ. Fed. Span. Oncol. Soc. Natl. Cancer Inst. Mex. 18, 160–171 (2016).

24. Sakaue, S. et al. A cross-population atlas of genetic associations for 220 human phenotypes. Nat. Genet. 53, 1415–1424 (2021).

25. Giannisis, A. et al. Plasma apolipoprotein E levels in longitudinally followed patients with mild cognitive impairment and Alzheimer’s disease. Alzheimers Res. Ther. 14, 115 (2022).

26. Rasmussen, K. L., Tybjærg-Hansen, A., Nordestgaard, B. G. & Frikke-Schmidt, R. Plasma levels of apolipoprotein E, APOE genotype, and all-cause and cause-specific mortality in 105 949 individuals from a white general population cohort. Eur. Heart J. 40, 2813–2824 (2019).

27. Fuior, E. V. & Gafencu, A. V. Apolipoprotein C1: Its Pleiotropic Effects in Lipid Metabolism and Beyond. Int. J. Mol. Sci. 20, 5939 (2019).

28. Yengo, L. et al. A saturated map of common genetic variants associated with human height. Nature 610, 704–712 (2022).

29. He, D. et al. A longitudinal genome-wide association study of bone mineral density mean and variability in the UK Biobank. Osteoporos. Int. J. Establ. Result Coop. Eur. Found. Osteoporos. Natl. Osteoporos. Found. USA 34, 1907–1916 (2023).

30. Chernus, J. et al. GWAS reveals loci associated with velopharyngeal dysfunction. Sci. Rep. 8, 8470 (2018).

31. Kurki, M. I. et al. FinnGen provides genetic insights from a well-phenotyped isolated population. Nature 613, 508–518 (2023).

32. Sudlow, C. et al. UK biobank: an open access resource for identifying the causes of a wide range of complex diseases of middle and old age. PLoS Med. 12, e1001779 (2015).

33. Pärn, K. et al. Genotype imputation workflow v3.0. (2018).

34. Bycroft, C. et al. The UK Biobank resource with deep phenotyping and genomic data. Nature 562, 203–209 (2018).

35. Zhou, W. et al. Efficiently controlling for case-control imbalance and sample relatedness in large-scale genetic association studies. Nat. Genet. 50, 1335–1341 (2018).

36. Loh, P.-R. et al. Efficient Bayesian mixed-model analysis increases association power in large cohorts. Nat. Genet. 47, 284–290 (2015).

37. Bulik-Sullivan, B. K. et al. LD Score regression distinguishes confounding from polygenicity in genome-wide association studies. Nat. Genet. 47, 291–295 (2015).

38. Dai, Y. et al. WebCSEA: web-based cell-type-specific enrichment analysis of genes. Nucleic Acids Res. 50, W782–W790 (2022).

39. Kanehisa, M., Furumichi, M., Sato, Y., Kawashima, M. & Ishiguro-Watanabe, M. KEGG for taxonomy-based analysis of pathways and genomes. Nucleic Acids Res. 51, D587–D592 (2023).

40. Milacic, M. et al. The Reactome Pathway Knowledgebase 2024. Nucleic Acids Res. 52, D672–D678 (2023).

41. Agrawal, A. et al. WikiPathways 2024: next generation pathway database. Nucleic Acids Res. 52, D679–D689 (2024).

42. McLaren, W. et al. The Ensembl Variant Effect Predictor. Genome Biol. 17, 122 (2016).

43. Sun, B. B. et al. Plasma proteomic associations with genetics and health in the UK Biobank. Nature 622, 329–338 (2023).

44. Ge, T., Chen, C.-Y., Ni, Y., Feng, Y.-C. A. & Smoller, J. W. Polygenic prediction via Bayesian regression and continuous shrinkage priors. Nat. Commun. 10, 1776 (2019).

45. Chou, W.-C. et al. A combined reference panel from the 1000 Genomes and UK10K projects improved rare variant imputation in European and Chinese samples. Sci. Rep. 6, 39313 (2016).

