## Supplementary information for "Large-scale genome-wide analyses with proteomics integration reveal novel loci and biological insights into frailty"

### Contents

Ethics statements of FinnGen and UK Biobank. 2

Supplementary Figure 1. Venn diagram showing the overlap between genes associated with the (a) HFRS and (b) HFRS without dementia at $p<5\times10^{-8}$ in FinnGen and those reported in the three previous frailty-related GWASs. 4

Supplementary Figure 2. QQ-plots for association summary statistics of (a) HFRS and (b) HFRS without dementia in FinnGen. 5

Supplementary Figure 3. Top 20 enriched cell types for HFRS in FinnGen. 6

Supplementary Figure 4. Top 20 enriched cell types for HFRS without dementia in FinnGen. 7

Supplementary Figure 5. Regional association plots for gene loci identified by colocalization analysis of cis-pQTL and HFRS GWAS 8

**Ethics statements of FinnGen and UK Biobank.**

*FinnGen*

Patients and control subjects in FinnGen provided informed consent for biobank research, based on the Finnish Biobank Act. Alternatively, separate research cohorts, collected prior the Finnish Biobank Act came into effect (in September 2013) and start of FinnGen (August 2017), were collected based on study-specific consents and later transferred to the Finnish biobanks after approval by Finnish Medicines Agency (Fimea), the National Supervisory Authority for Welfare and Health. Recruitment protocols followed the biobank protocols approved by Fimea. The Coordinating Ethics Committee of the Hospital District of Helsinki and Uusimaa (HUS) statement number for the FinnGen study is Nr HUS/990/2017. The FinnGen study is approved by Finnish Institute for Health and Welfare (permit numbers: THL/2031/6.02.00/2017, THL/1101/5.05.00/2017, THL/341/6.02.00/2018, THL/2222/6.02.00/2018, THL/283/6.02.00/2019, THL/1721/5.05.00/2019 and THL/1524/5.05.00/2020), Digital and population data service agency (permit numbers: VRK43431/2017-3, VRK/6909/2018-3, VRK/4415/2019-3), the Social Insurance Institution (permit numbers: KELA 58/522/2017, KELA 131/522/2018, KELA 70/522/2019, KELA 98/522/2019, KELA 134/522/2019, KELA 138/522/2019, KELA 2/522/2020, KELA 16/522/2020), Findata permit numbers THL/2364/14.02/2020, THL/4055/14.06.00/2020,,THL/3433/14.06.00/2020, THL/4432/14.06/2020, THL/5189/14.06/2020, THL/5894/14.06.00/2020, THL/6619/14.06.00/2020, THL/209/14.06.00/2021, THL/688/14.06.00/2021, THL/1284/14.06.00/2021, THL/1965/14.06.00/2021, THL/5546/14.02.00/2020, THL/2658/14.06.00/2021, THL/4235/14.06.00/202, Statistics Finland (permit numbers: TK-53-1041-17 and TK/143/07.03.00/2020 (earlier TK-53-90-20) TK/1735/07.03.00/2021, TK/3112/07.03.00/2021) and Finnish Registry for Kidney Diseases permission/extract from the meeting minutes on 4th July 2019.

The Biobank Access Decisions for FinnGen samples and data utilized in FinnGen Data Freeze 9 include: THL Biobank BB2017_55, BB2017_111, BB2018_19, BB_2018_34, BB_2018_67, BB2018_71, BB2019_7, BB2019_8, BB2019_26, BB2020_1, Finnish Red Cross Blood Service Biobank 7.12.2017, Helsinki Biobank HUS/359/2017, HUS/248/2020, Auria Biobank AB17-5154 and amendment #1 (August 17 2020), AB20-5926 and amendment #1 (April

23 2020) and it´s modification (Sep 22 2021), Biobank Borealis of Northern Finland_2017_1013, Biobank of Eastern Finland 1186/2018 and amendment 22 § /2020, Finnish Clinical Biobank Tampere MH0004 and amendments (21.02.2020 & 06.10.2020), Central Finland Biobank 1-2017, and Terveystalo Biobank STB 2018001 and amendment 25th Aug 2020.

*UK Biobank*

The UK Biobank study was approved by the North West Multi-Centre Research Ethics Committee (approval number: 11/NW/03820). All participants provided written informed consent for data collection, analysis, and record linkage. We have also obtained an ethical approval for the use of UK Biobank data in Sweden (Dnr 2016/1888-31/1).

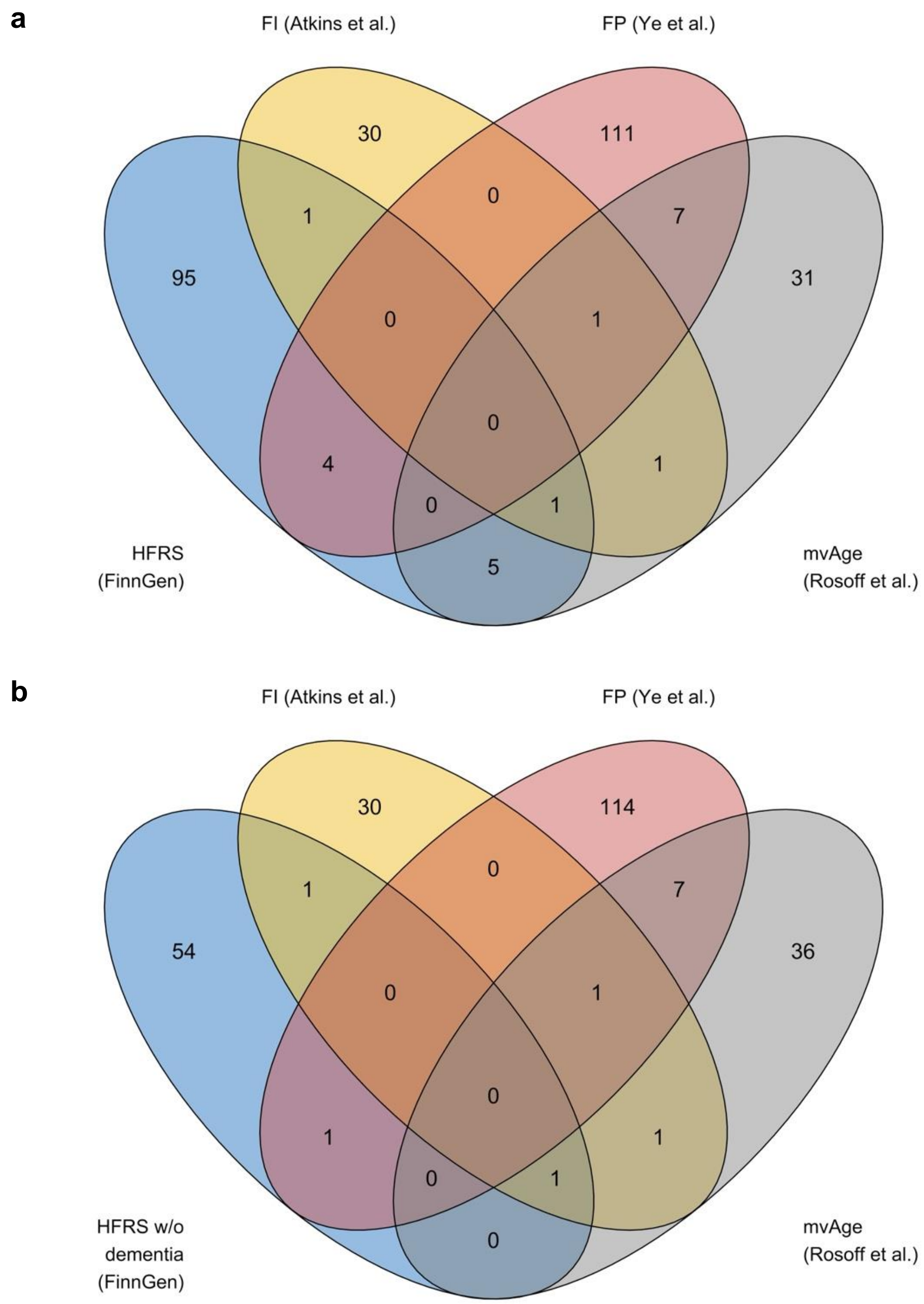


**Supplementary Figure 1.** Venn diagram showing the overlap between genes associated with the **(a)** HFRS and **(b)** HFRS without dementia at $p<5\times10^{-8}$ in FinnGen and those reported in the three previous frailty-related GWASs.

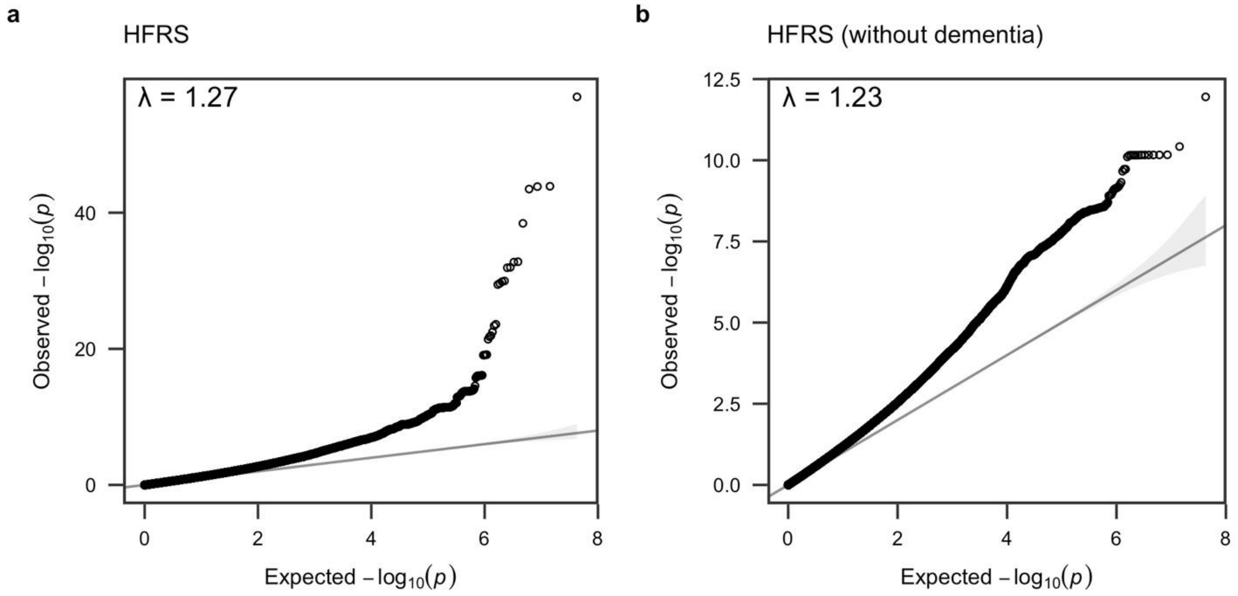


**Supplementary Figure 2.** QQ-plots for association summary statistics of **(a)** HFRS and **(b)** HFRS without dementia in FinnGen.

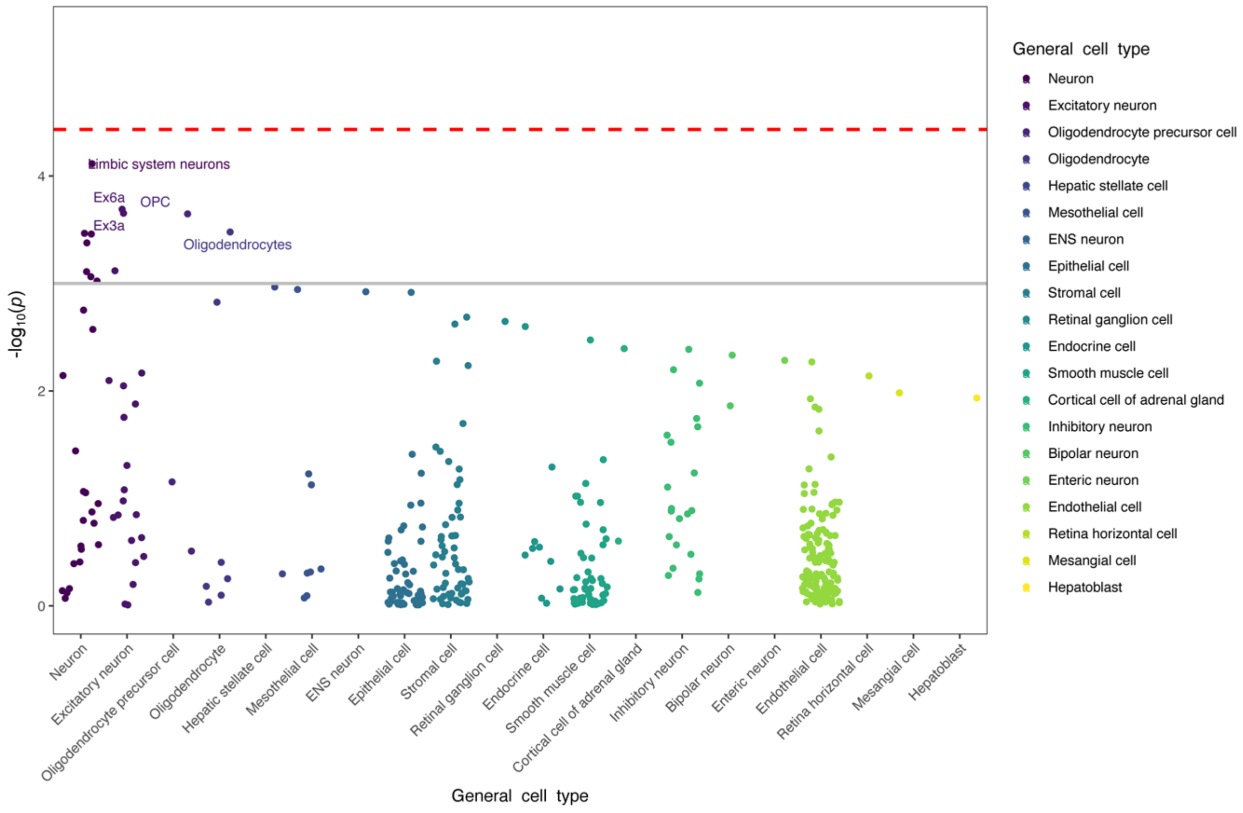


**Supplementary Figure 3.** Top 20 enriched cell types for HFRS in FinnGen.

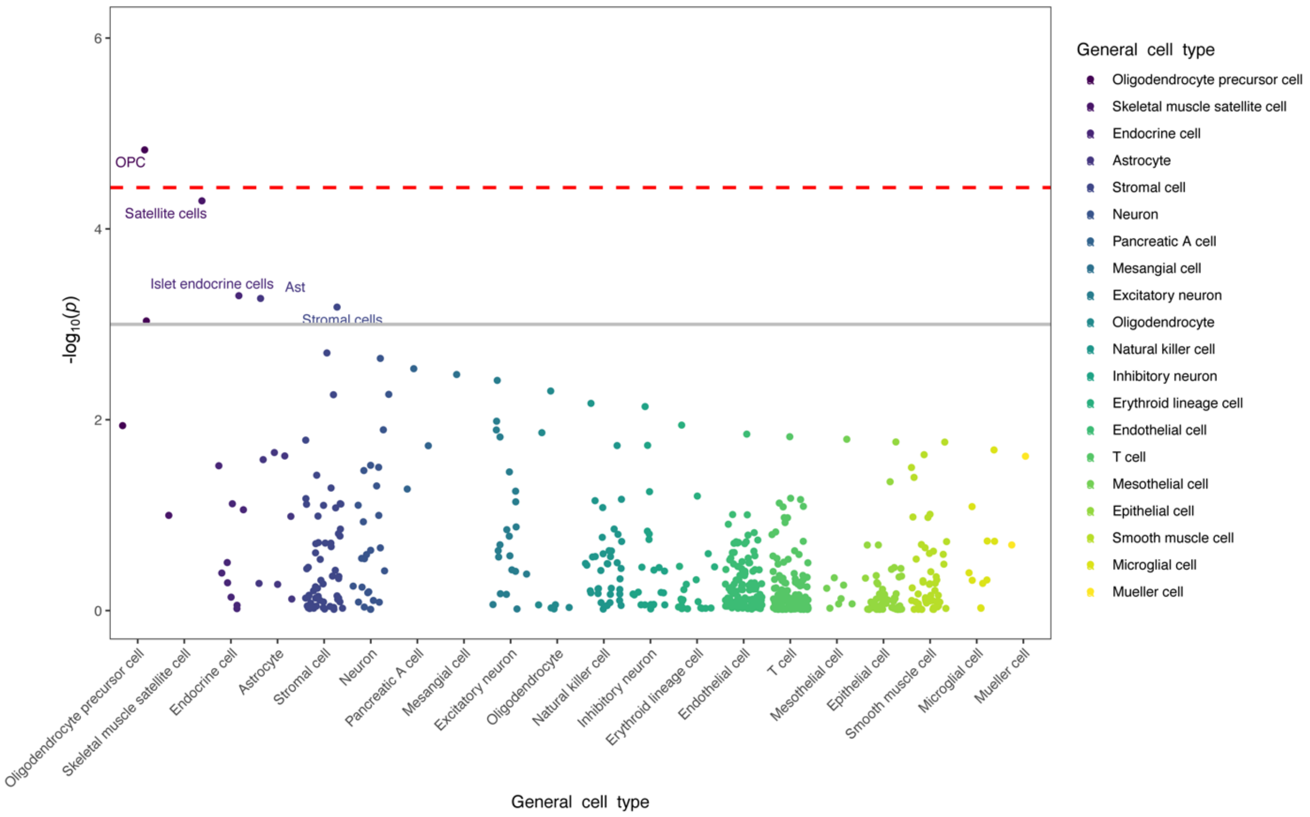


**Supplementary Figure 4.** Top 20 enriched cell types for HFRS without dementia in FinnGen.

A.

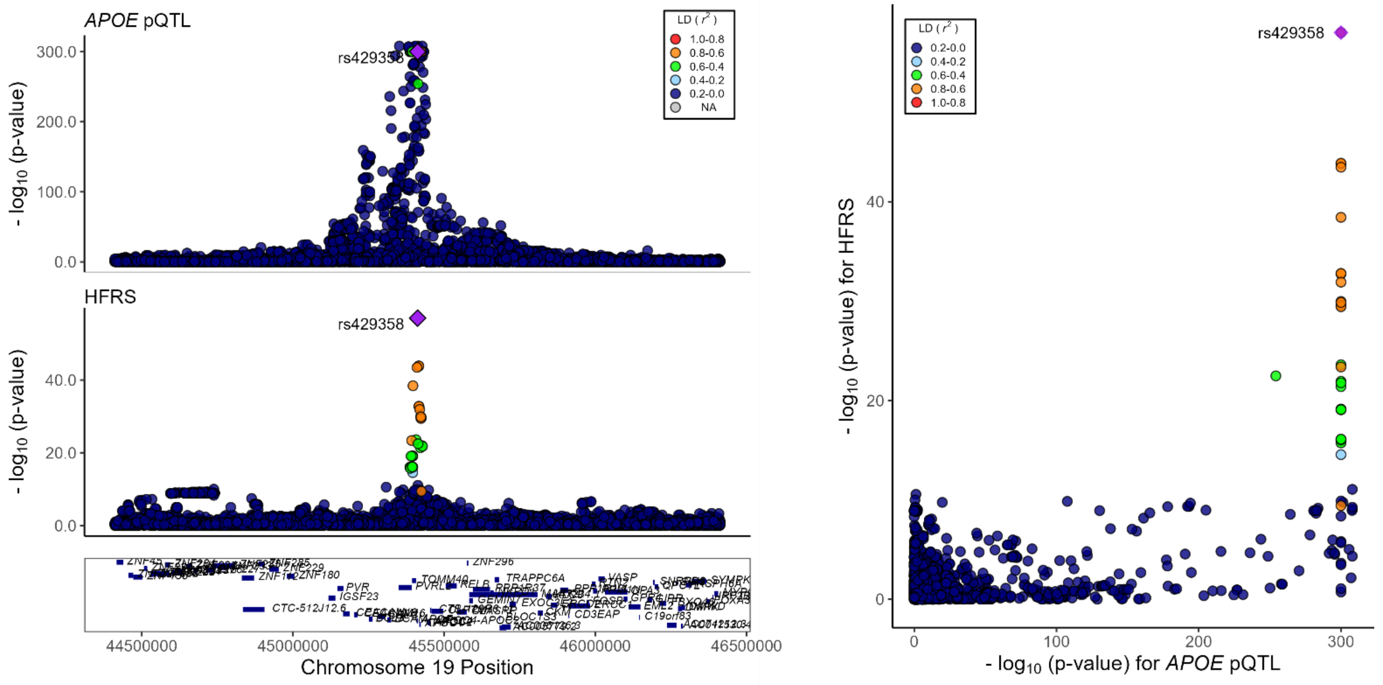


B.

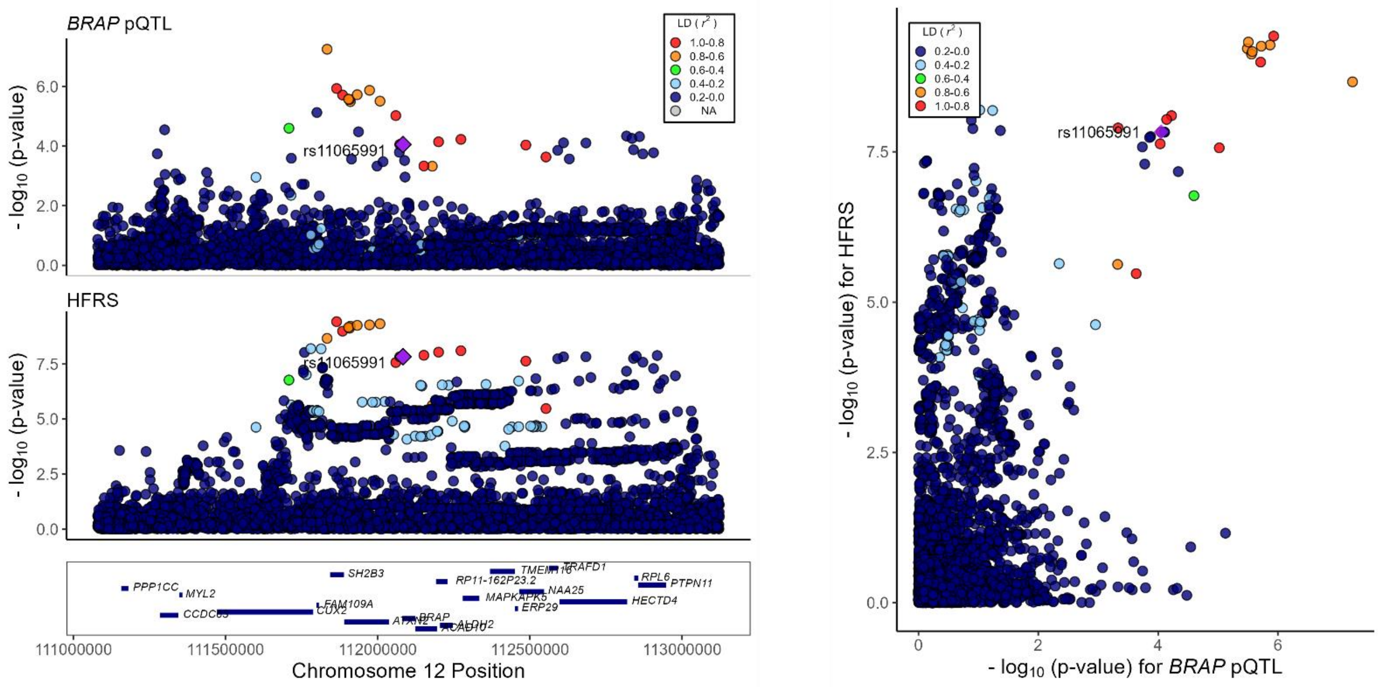


**Supplementary Figure 5.** Regional association plots for gene loci identified by colocalization analysis of cis-pQTL and HFRS GWAS.
